# Human *ace2* and *tmprss2* polymorphisms for predicting susceptibility to tuberculosis and COVID-19 co-infection in Cameroonian cohort

**DOI:** 10.1101/2024.11.14.24317326

**Authors:** Mary Ngongang Kameni, Eric Berenger Tchoupe, Severin Donald Kamdem, Nikhil Bhalla, Assam Assam Jean Paul, Tepa Njiguet Arnaud, Fuh Roger Neba, Ranjan Kumar Nanda, Anthony Afum-Adjei Awuah, John Amuasi, Palmer Masumbe Netongo

## Abstract

SARS-CoV-2 and *Mycobacterium tuberculosis* (*Mtb*) share similarities in their modes of transmission, pathophysiological symptoms, and manifestations. An imbalance in the immune response characterized by significantly elevated levels of some inflammatory cytokines may increase the risk of developing both tuberculosis (TB) and COVID-19 as a comorbid condition. The role of SNPs in *ace2* and *tmprss2* conferring higher susceptibility to TB-COVID-19 co-infection is relatively underexplored. In this study, a Cameroonian cohort consisting of COVID-19-infected (n = 31), TB-infected (n = 43), TB-COVID-19 co-infected (n = 21), and a control group (n = 24) was studied. The immune response and disease severity were estimated by quantitating inflammatory cytokine levels and self-reported and clinically diagnosed symptoms. We identified SNPs in *ace2* and *tmprss2* genes previously associated with COVID-19 susceptibility and assessed their association with comorbid conditions. We identified genotypes (Allele AG: rs147311723, rs35803318; Allele AA: rs2074192; Allele CG: rs4240157; Allele AG: rs4646179) in *ace2* gene and (Allele CA: rs61735791, Allele CT: rs12329760) in *tmprss2* genes that are putatively associated with higher susceptibility to both TB and COVID-19. This study underscores the significant genetic and immunological factors contributing to susceptibility to TB and COVID-19 co-infections.

## Introduction

Tuberculosis (TB) is a global public health concern and is endemic in low to middle-income countries, mostly those in sub-Saharan Africa and Asia. According to the Global TB Report 2023 by the World Health Organisation (WHO), 8.2 million new cases and 1.25 million deaths were reported in 2023 alone, and TB was the topmost cause of mortality and morbidity before the COVID-19 pandemic (1). The COVID-19 pandemic overtook TB-related deaths and incidences, making it the most infectious disease with the highest mortality after April 2020. COVID-19, first reported in Hubei Province in China in December 2019, is caused by Severe Acute Respiratory Disease Syndrome Coronavirus 2 (SARS-CoV-2) (2). During the same period, 3,931,534 cases and 100,952 deaths due to COVID-19 were reported in sub-Saharan Africa (SSA) (6). SARS-CoV-2 and *Mycobacterium tuberculosis* (Mtb) show similar modes of transmission, pathophysiological symptoms and manifestations. They transmit through respiratory tract secretions via aerosol mode and cause lower respiratory tract infections, pneumonia and associated lung fibrosis. Both diseases share similar clinical symptoms, such as cough, fever, weakness, and dyspnea (7). An imbalance of immune response, including significantly decreased absolute counts of T-cells and increased pro-inflammatory cytokines levels, may influence the risk of developing TB and COVID-19 comorbid conditions, among others (8). The cytokine storm causes a major biological response identified in patients with severe COVID-19 due to the over-activation of the inflammatory cascade in the tissues exposed to harmful stimuli like injury, toxic chemicals or pathogens. (9).

The angiotensin-converting enzyme 2 (*ace2*) gene is located on the long arm of human chromosome 17 and consists of 26 exons and 25 introns (17q23.3) with a mass of ∼92.5 kDa (10). Ace2 catalyses the conversion of angiotensin I into angiotensin 1-9, and angiotensin II into the vasodilator angiotensin 1-7 (11). Ace2 possesses a catalytic and collectrin-like domain that spans the membrane and makes it a surface protein. The spike protein of SARS-COV-2 binds to the catalytic domain of Ace2, leading to a conformational change, thereby causing the viral entry into the host cell (12). Transmembrane protease serine 2 (Tmprss2) is a protein encoded by the *tmprss2* gene, located on autosomal chromosome 21q22.3, which regulates cell signaling and modulates host response to infection (13). However, in SARS-COV-2 infection, *tmprss2* primes the viral glycoprotein by leaving the spike protein at the S1/S2 junction and the S2 site (3). Both *tmprss2* and *ace2,* expressed in the lung bronchial epithelial cells, are therefore critical for viral entry of SARS-COV-2 (4). Some studies have demonstrated that *tmprss2* activity is essential for viral spread and pathogenesis, enabling the entry of SARS-CoV-2 in *ace2*-expressing cells but not in cells without *ace2* (14,15). Ziegler and colleagues had shown that *Mtb* infection increases *ace2* expression in the lung tissues through interferon stimulation (16). Susceptibility to SARS-COV-2 infection could be defined by the age, gender and pre-existing comorbidities like health status, genetic background, immunological state of the host and also depends on the lineage of the pathogen (17). The host *ace2* and *tmprss2* genes play a vital role in disease severity, viral replication, and inflammation, and variations at genetic levels, such as single nucleotide polymorphisms (SNPs), may affect their function (5). The SNPs distribution in these genes could be population and/or clade-specific and may contribute to higher susceptibility to COVID-19 or TB. Limited literature is present on the effect of SNPs in *ace2* and *tmprss2* and their response to COVID-19 and TB comorbid conditions. In this study, 119 subjects with TB, COVID-19 or both were recruited from Cameroon; their biochemical and immunological parameters and qRT-PCR-based genotyping assays were carried out to determine the associated mutation pattern in *ace2* and *tmprss2* shedding light on comorbidity susceptibility and SNPs.

## Results

### Socio-demographic and clinical characteristics of study participants

119 participants from the Central region of Cameroon were included in this study. All study participants presented signs and symptoms of a respiratory disease. Participants were enrolled at the Djoungolo District Hospital (102), Jamot Hospital (n=83), Ekoumdoum Baptist Hospital (n=118) and Red Cross Hospital (n=96) in Yaounde during the COVID-19 pandemic from the period of March 2021 to September 2022 (Figure 1A). Every participant underwent clinical examination and laboratory assessment for COVID-19 and TB. Recruited participants were assessed for medical history, tobacco consumption and TB medication history. The study populations consisted of four groups: COVID-19 (n= 31), TB (n= 43), TB-COVID-19 co-infection (n= 21) and a set of healthy controls (n= 24) (Figure 1 B). The proportion of males to females was 60% and 40%, respectively. The most frequent symptoms in the COVID-19 group (n=31) were cough (74%), fever (70.9%) and headache (61.3%). The main clinical signs reported during COVID-19/TB co-infection (n=21) were cough (85.7 %), fatigue (80.9%) and headache (76.1 %). TB patients (n=43) presented predominant symptoms such as cough (53.5 %), fever (46.6 %) and chest pain (25.6 %). Details of sociodemographic and clinical data are summarized in (Table 1). Patient recruitment workflow is illustrated in the form of a flowchart in Figure 1B.

**Figure 1:**
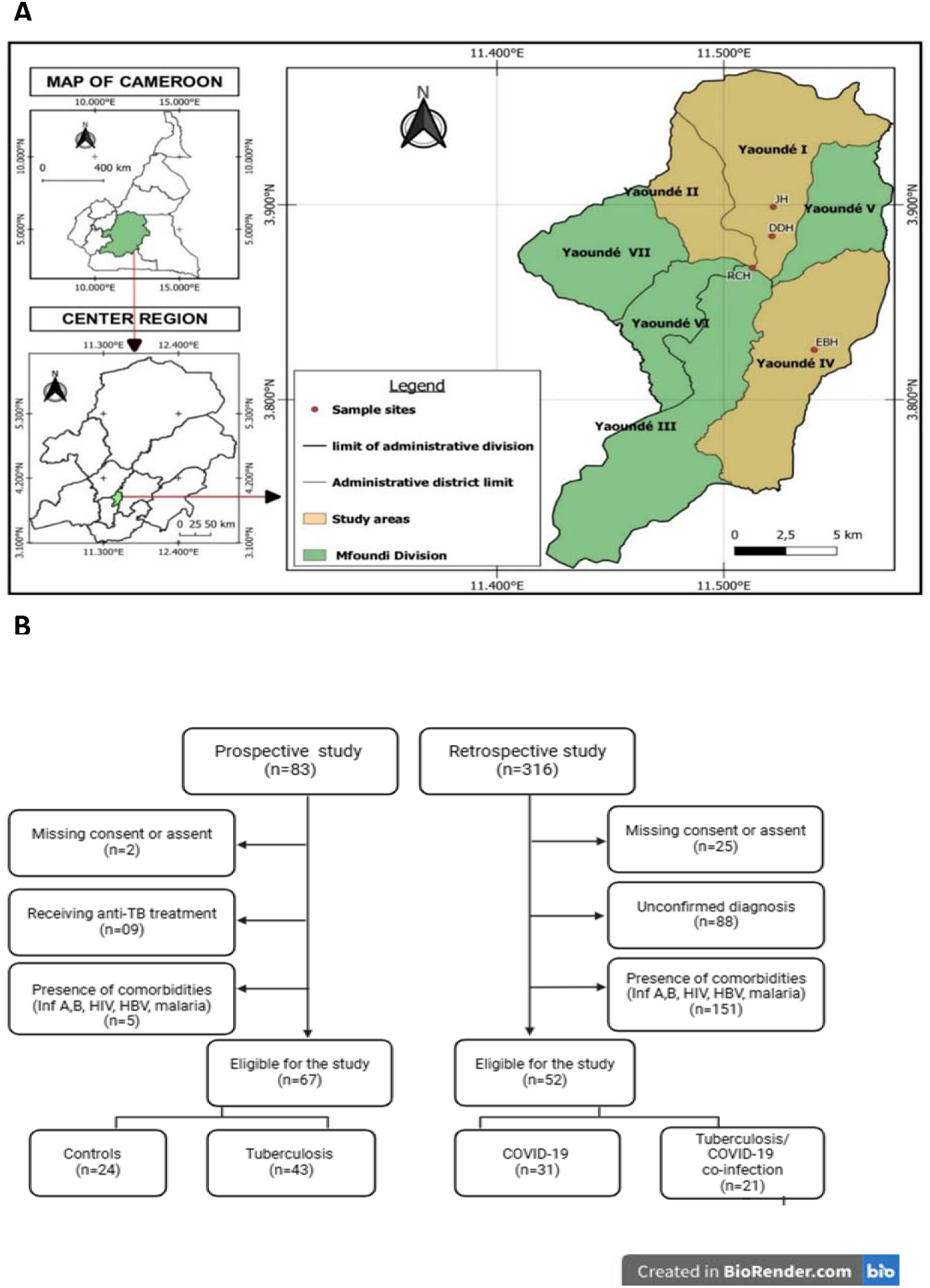
Study participant’s recruitment and classification. A: Map showing the Centre region of Cameroon with different study sites. Samples for this study were collected at 4 hospitals: JH, DDH, RCH, and EBH, which are situated in Yaoundé. The sample sizes from each district were as follows: JH: 83; DDH: 102; RCH: 96; and EBH: 118. The map was created using QGIS version 3.32.3. B: Based on COVID-19 and Tuberculosis infection status, participants were grouped into 4 groups: Controls, COVID-19 positive, Tuberculosis positive and Tuberculosis/COVID-19 co-infected. In total, our sample size consisted of 119 participants. Abbreviations: JH: Jamot Hospital, DDH: Djoungolo District Hospital, RCH: Red Cross Hospital, EBH: Ekoumdoum Baptist Hospital.

**Table 1:** Socio-demographic and clinical characteristics of study participants from the four hospitals situated in the centre region of Cameroon. The vitals were measured by clinicians at the time of patient enrolment using thermometer for temperature; sphygmomanometer for heart rate and blood pressure;. Other symptoms viz, Cough, Sore throat, Bloody sputum, Rhinorrhoea, Chest pain, Myalgia, Arthralgia, Fatigue, Loss of smell and headache were self-reported by participants and noted by clinicians at the time of enrolment. Abbreviations: SD: standard deviation; ns: non significant; bpm: beats per minutes; bm; breathes per minutes °C: degree celcius; O2: oxygen; COV: COVID-19; TB: Tuberculosis; TBCOV: Tuberculosis-COVID-19 co-infection.

| Characteristics | Control (24) | COV (31) | TB (43) | TBCOV (21) | P-values |
| --- | --- | --- | --- | --- | --- |
| Age (mean ± SD) | 30.5±6.7 | 37.3±12.8 | 36.8±17.2 | 37.7±14.5 | ns |
| Male n (%) | 11 (45) | 17 (55) | 32 (74) | 13 (62) | N/A |
| Female n (%) | 13 (55) | 14 (45) | 11 (36) | 8 (38) | N/A |
| Heart Rate bpm (mean ± SD) | 88.5±16.4 | 87.5±10.6 | 86.6±10.5 | 87.5±8.7 | ns |
| Respiratory Rate bm (mean ±SD) | 18.9±1.3 | 16.8±2.8 | 17.3±3.1 | 17.9±1.2 | p<0.05 |
| Systolic blood pressure BP/mmHg (mean ± SD) | 117.8±10.9 | 122.9±19.0 | 125.3±20.1 | 118.5±8.4 | ns |
| Diastolic Blood Pressure BP/mmHg (mean ± SD) | 76.9±8.8 | 80.7±14.7 | 77.8±13.8 | 79.8±10.2 | ns |
| O2 saturation % (mean ± SD) | 98.0±1.3 | 96.6±1.8 | 97.9±1.1 | 95.7±3.2 | p<0.05 |
| Fever n (%) | 11 (45.5) | 22 (70.9) | 20 (46.6) | 15 (71.4) | P=0.05 |
| Cough n (%) | 12 (50) | 23 (74.2) | 23 (53.5) | 18 (85.7) | ns |
| Sore throat n (%) | 5 (22.7) | 9 (29) | 10 (23.3) | 9 (45) | ns |
| Bloody sputum n (%) | 0 (0) | 1 (3.2) | 7 (16.3) | 2 (10) | p<0.05 |
| Rhinorrhoea n (%) | 10 (41.7) | 12 (38.7) | 7 (16.3) | 7 (35) | ns |
| Chest pain n (%) | 5 (22.7) | 9 (29) | 11 (25.6) | 12 (57.1) | ns |
| Myalgia n (%) | 5 (22.7) | 12 (38.7) | 9 (20.9) | 8 (40) | ns |
| Arthralgia n (%) | 12 (50) | 6 (19.4) | 9 (20.9) | 11 (52.3) | ns |
| Fatigue n (%) | 12 (50) | 13 (41.9) | 9 (20.9) | 17 (80.9) | p<0.05 |
| Loss of smell n (%) | 5 (20.8) | 9 (29.0) | 6 (13.9) | 6 (30) | ns |
| Headache n (%) | 11 (45.8) | 19 (61.3) | 7 (16.3) | 16 (76.1) | p<0.05 |

\*\*\*End of the Manuscript\*\*\*

### Serum AST, Urea and D-dimer levels are significantly higher in the Tuberculosis and COVID-19 comorbid group

The blood clotting factor and other biochemical parameters of every study participant were evaluated in order to monitor their liver and kidney function (Figure 2A). TB-COVID-19 co-infected patients had higher AST and Urea levels than individual TB and COVID-19 patients. AST levels were higher than the normal range (> 34 IU/L) in 50% of the TB-COVID-19 co-infected group, 40 % of TB and 29% of COVID-19 participants (Figure 2A). It was noticed that every TB and COVID-19 patient had urea levels exceeding the normal serum concentration (> 20 mg/dL). Conversely, 82% of COVID-19 and 93% of TB-positive patients exhibited abnormal urea levels (Figure 2A). The COVID-19 group exhibited significantly elevated ALT levels (p < 0.001) compared to both the TB and TB-COVID-19 groups. ALT levels exceeding 36 IU/L were found in 29% of COVID-19 patients, and 15% of TB-COVID-19 co-infected patients (Figure 2A). The D-dimer levels, which are essential for evaluating coagulation abnormalities in clinical settings, were measured among the study participants. In this study, over 60% of TB and TB-COVID-19 patients demonstrated elevated D-dimer levels (> 0.5 mg/dL) compared to just 23% of control subjects. Notably, a small subset of COVID-19 patients also exhibited abnormal D-dimer levels, highlighting the test’s significance in this context. Serum creatinine levels were significantly higher in TB patients than those with COVID-19 or TB-COVID-19 co-infection (Figure 2A). Elevated creatinine levels exceeding 1.4 mg/dL were noted in some of the patients, 18% of those with COVID-19, 24% of TB patients, and 20% of those co-infected with TB and COVID-19 showing abnormally high values. Direct bilirubin and total bilirubin levels were strikingly similar across all four groups, highlighting a consistent biochemical profile. Total serum bilirubin levels were abnormal (> 1.2 mg/dL) in three subgroups: 9% of COVID-19 patients, 37% of TB patients, and 45% of the TB-COVID-19 co-infection group (Figure 2A). Furthermore, direct bilirubin levels were elevated (> 0.5 mg/dL) in the majority of TB (37%) and TB-COVID-19 (45%) patients, in contrast to just 4% of COVID-19 patients and 23% of healthy controls. (Figure 2A).

**Figure 2:**
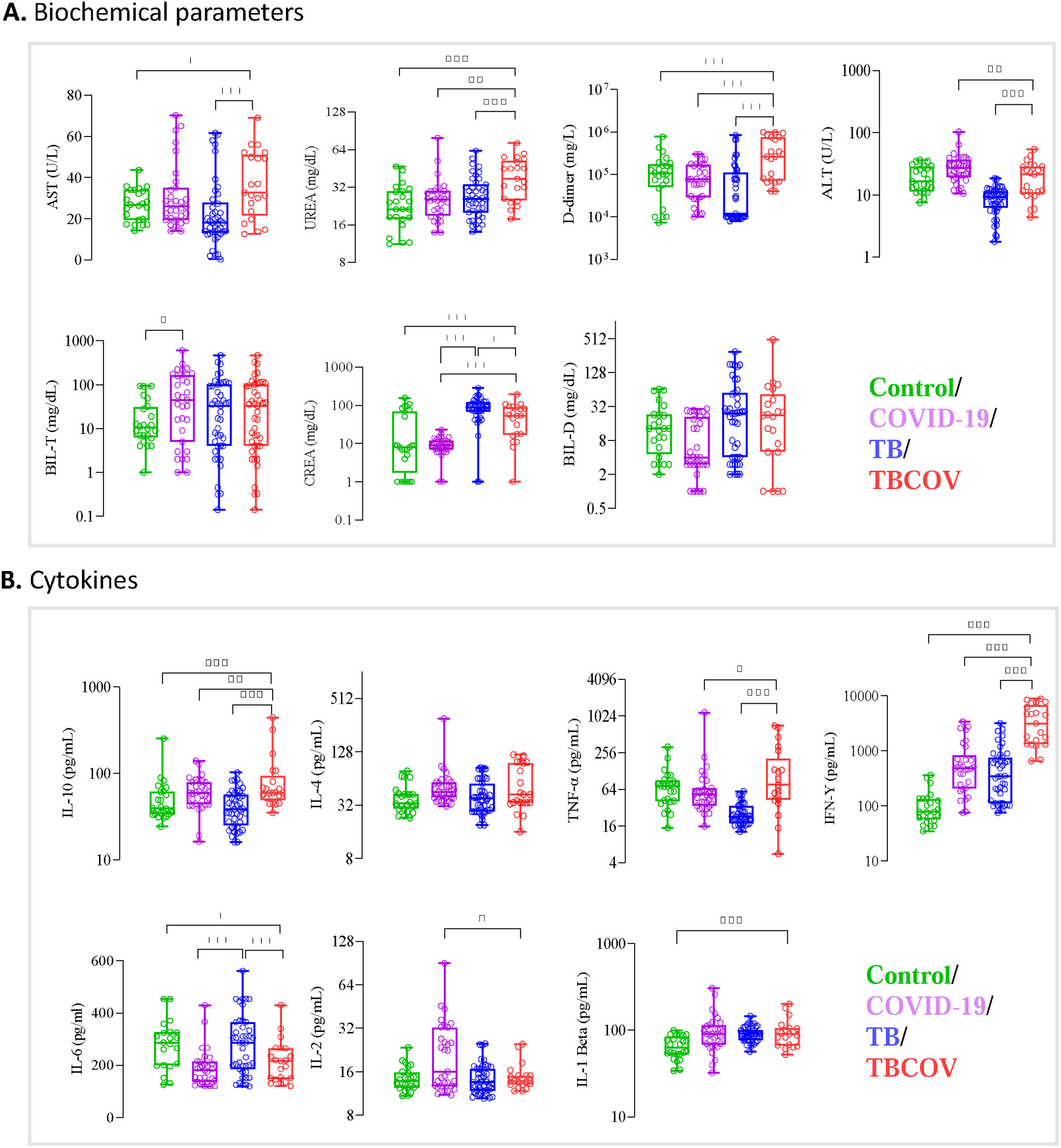
**Increased serum levels of AST, UREA, D-dimer and plasma levels of IL-4, IL-10, TNF-**α**, IFN-**γ **showed association with Tuberculosis and COVID-19 co-infection.** Each individual’s data point is shown, with median values indicated by horizontal lines. A: Biochemical parameters such as AST, ALT, D-dimer, Total-bilirubin, Direct-bilirubin, Urea and Crea were measured using spectrophotometry-based assays. B: Plasma levels of anti-inflammatory cytokines are associated with TB/COVID-19 co-infection. Sandwich ELISA using Origene kits was employed to measure the systemic levels of anti-inflammatory cytokines (IL-10 and IL-4) and pro-inflammatory (TNF-α, IFN-γ, IL-6, IL-2 and IL-β) in plasma samples of the study population. Unpaired t-test was carried out between various combinations to determine statistically significant differences. Abbreviations: AST: Aspartate aminotransferase; ALT: Alanine aminotransferase; CREA: Creatinine; TNF: Tumour necrosis factor; INF: Interferon; IL: interleukin; TB: tuberculosis; COV: Covid-19; TBCOV: Tuberculosis and Covid-19 co-infection. *p <0.05, **p <0.01, ***p <0.001.

### Higher plasma IL10, IFN-γ and TNF-α levels are associated with TB-COVID-19 co-infections

In order to monitor the inflammation score, plasma IFN-γ, TNF-α, IL-6, IL-10, IL-4, IL-2 and IL-1β levels were monitored between study groups. Higher plasma levels of two pro-inflammatory cytokines (IFN-γ, TNF-α) were found to be associated with TB-COVID-19 co-infection. Plasma INF-γ levels were significantly higher in the TB-COVID-19 co-infected subjects than in COVID-19 or TB co-infected patients. Plasma TNF-α were significantly higher in TB-COVID-19 co-infected patients than in either TB or COVID-19 mono-infection cases. In COVID-19 subjects, plasma IL-2 levels were significantly higher compared to TB-COVID-19 and TB patients. Plasma IL-1β levels did not vary across the diseased groups (COVID-19, TB and TB-COVID-19) but were significantly lower in control participants. TB-positive patients expressed significantly higher plasma IL-6 levels compared to TB-COVID-19 and COVID-19-positive patients. Among the two anti-inflammatory cytokines that were measured in this study, plasma IL-10 levels were significantly higher in TB-COVID-19 co-infected patients compared to the TB (p<0.0001) and COVID-19 (p<0.01) patients. Plasma IL-4 levels expressed by COVID-19, TB and TB-COVID-19 patients did not vary significantly. (Figure 2B).

### Variations in SNPs of *ace2* and *tmprss2* gene are associated with disease susceptibility to COVID-19 and Tuberculosis mono- and co-infections

The SNPs positions in *ace2* (rs2285666, rs4240157, rs4646142, rs4646116, rs6632677, rs4646140, rs147311723, rs2074192, rs35803318, rs4646179) and *tmprss2* (rs12329760, rs75603675, rs61735791) were profiled in every study participant (Figure 3A and 3B). Analysis revealed a significant statistical association among the four groups concerning genotypes for five SNPs, including rs4646142 (p < 0.05), rs2074192 (p < 0.05), rs147311723 (p < 0.0001), rs4646140 (p < 0.05), and rs2285666 (p < 0.001). The principal findings of our study, detailed in Supplementary Table 1, highlighted an increase in heterozygous genotype frequencies for *ace2* rs4646140 and rs35803318 (AG), as well as *tmprss2* rs61735791 (CA) in the TB/COVID-19 co-morbid group. In this study, we observed a significant increase in the genotype frequencies of *ace2* rs4646116, rs2285666, and rs4646179 (AG, CA, and AG, respectively) in COVID-19. Additionally, the heterozygous genotype frequencies of *ace2* rs4646142, rs35803318, rs4646116, rs2285666 (AG), and *tmprss2* rs75603675 and rs61735791 (CA) were notably prevalent in TB patients. Homozygous/double mutant alleles of ACE2 rs2074192 (AA) were frequently observed in TB patients (n=32) and those co-infected with TB and COVID-19 (n=17) (Figure 3). In addition, heterozygous mutant alleles of *tmprss2* rs147311723 (AA) were particularly prevalent among TB patients. Among TB patients, the most common genotypes included AG for rs4646142 and rs4646116, as well as CA for rs2285666, rs75603675, and rs61735791. The wild-type alleles of the *ace2* gene, specifically rs4646140 (GG), were found to be more prevalent in the disease groups compared to controls. Among COVID patients, rs2285666 was the most frequent SNP in 100% of patients exhibiting the CA genotype. Additionally, the second most frequent SNPs in this group were rs4646116 and rs4646179, where 97 % of patients had the AG genotype and 4 % had the GG genotype. While there was variation in SNP frequencies across the study population, SNP rs61735791 showed a notable association with TB and TB/COVID-19 co-infection.

**Figure 3:**
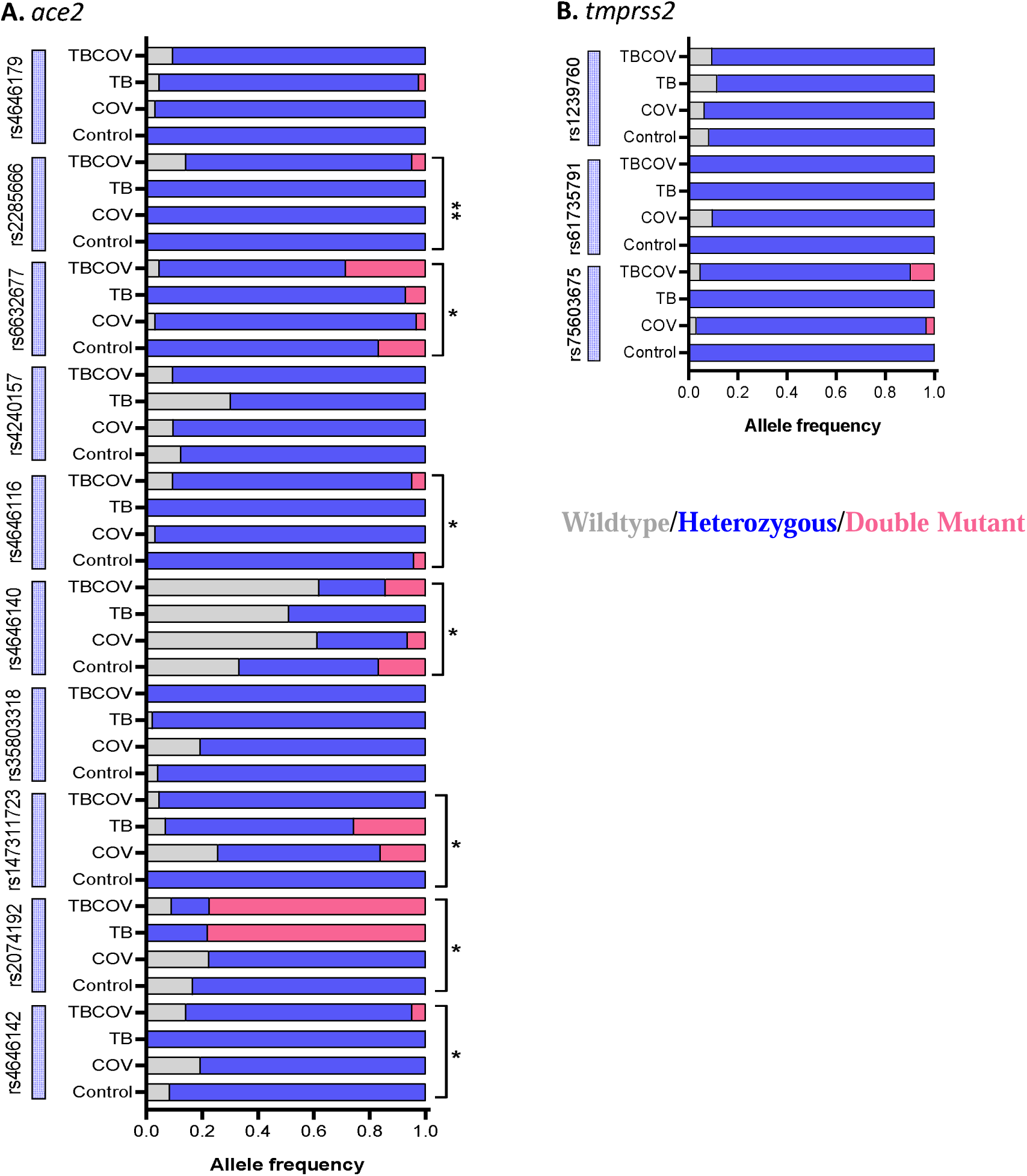
Double mutant *ace2* rs2074192, rs6632677 and *tmprss2* rs75603675 genotypes are associated with susceptibility to Tuberculosis and COVID-19 co-infection: Genomic DNA isolated using Zymo Human DNA Isolation Kit was subjected to quantitative real-time PCR with TaqMan probes. TaqMan SNP Genotyping Assays were employed to identify SNPs in *ace2* and *tmprss2* genes. A: specific genotype frequencies in *ace2.* B: specific genotype frequencies in *tmprss2*. Multigroup analysis was carried out using the Chi-square test, and SNPs having a p-value of less than 0.05 were considered to have a significantly perturbed presence between the groups (Control, TB, COV, and TBCOV). Abbreviations: TB: Tuberculosis positive; COV: COVID-19 positive and TBCOV: Tuberculosis and COVID-19 co-infection.

### Correlations between SNPs, Biochemical Markers and Cytokine Profiles

In this study, we investigated the potential correlations between the prevalence of identified *ace2* and *tmprss2* SNPs and the levels of cytokines and biochemical markers across four groups: COVID-19 patients, TB patients, TB-COVID-19 co-infected patients, and controls. In healthy controls, mild correlations were observed between the investigated biomarkers. Haemoglobin levels were positively correlated with hematocrit levels. AST levels showed a negative correlation with total bilirubin, while D-dimer levels were positively correlated with creatinine levels. TNF-α exhibited a negative correlation with lymphocyte count, and IL-1β was positively correlated with both platelet and creatinine counts. IFN-γ showed a negative correlation with WBC count. Several SNPs in the *ace2* and *tmprss2* genes also demonstrated significant correlations in healthy controls. Specifically, rs12329760 was negatively correlated with IL-10 levels, rs6632677 showed a positive correlation with TNF-α, and rs4646116 was positively correlated with total bilirubin levels (Figure 4A).

**Figure 4:**
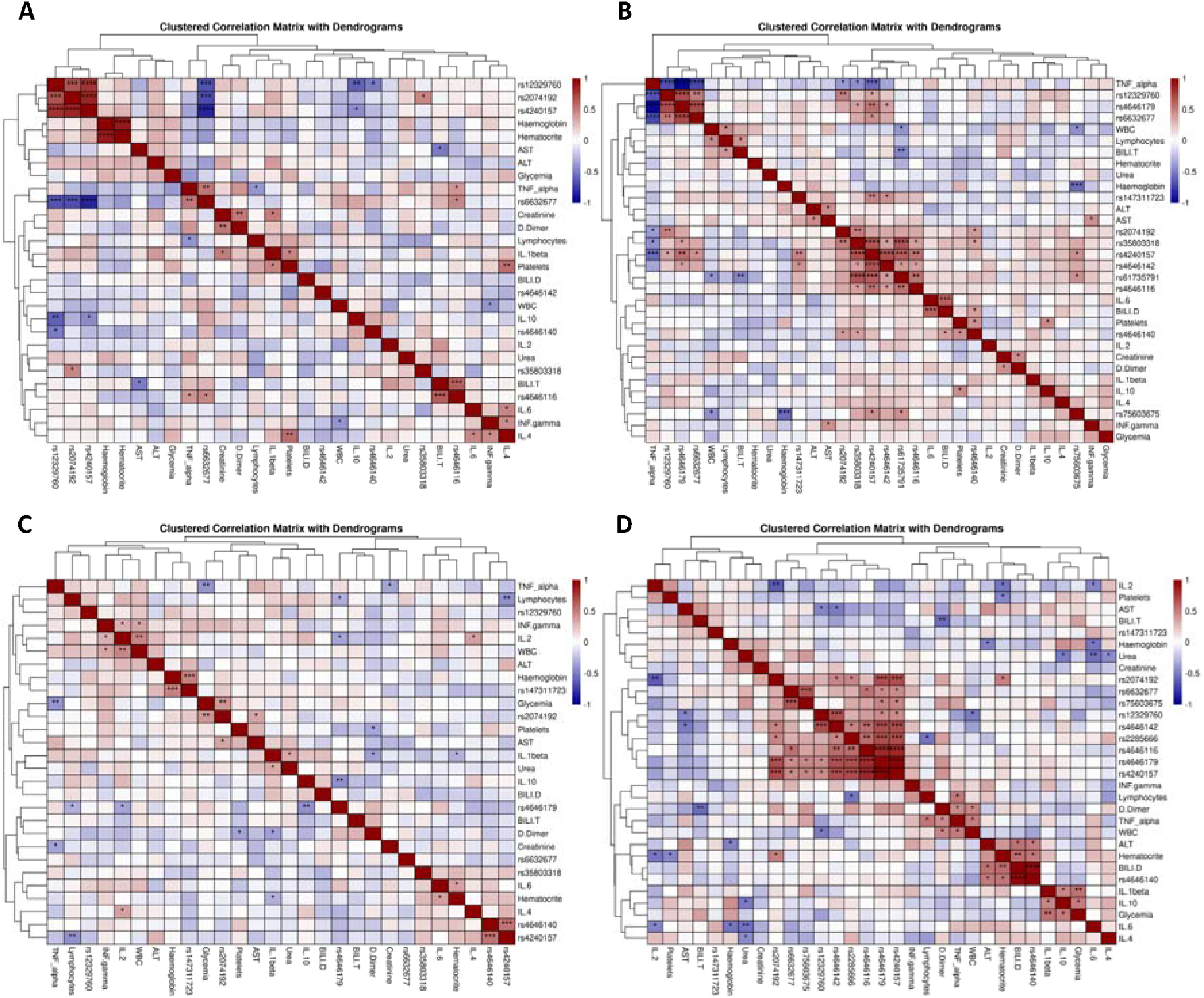
Correlation matrix between biochemical parameters, cytokine levels, and SNP profiles. The values were normalized to their respective Z-scores before the computation of correlation scores. A: Healthy; B: COV; C: TB and D: TBCOV. P-values: <0.05*; <0.01**; <0.001***; <0.0001****. Abbreviations: COV: COVID-19 group, TB: Tuberculosis infected group, and TBCOV: Tuberculosis and COVID-19 coinfected group. Note: Supplementary Figure is provided as a separate file.

In COVID-19 patients, several key biochemical markers showed significant correlations. AST levels exhibited a strong positive correlation with ALT. Creatinine levels were also positively correlated with D-dimer. Furthermore, AST levels were significantly positively correlated with IFN-γ, while direct bilirubin levels were positively correlated with IL-6, IL-10, and platelet count. Several *ace2* gene variants were significantly associated with cytokine levels. Specifically, the SNPs rs4240157, rs35803318, rs2074192, rs4646179, and rs66322677 in the *ace2* gene demonstrated significant negative correlations with TNF-α. Additionally, rs61735791 was negatively correlated with total bilirubin and WBC count. TMPRSS2 gene SNPs also exhibited significant correlations, with rs12329760 showing a negative correlation with TNF-α and rs75603675 being negatively correlated with both haemoglobin levels and WBC count (Figure 4B).

In TB patients, D-dimer levels were negatively correlated with both platelet count and IL-1β levels. Urea levels were positively correlated with IL-1β, while TNF-α exhibited a negative correlation with glucose levels. IL-2 levels were positively correlated with both WBC count and IFN-γ, whereas creatinine levels were negatively correlated with TNF-α. IL-6 exhibited a positive correlation with hematocrit levels. SNPs in the *ace2* and *tmprss2* genes were also significantly associated with biochemical markers in TB patients. Rs147311723 was positively correlated with haemoglobin levels, rs2074192 was positively correlated with both AST and glucose levels, rs4646179 showed a negative correlation with both IL-10 and IL-2, and rs4240157 was negatively correlated with lymphocyte count (Figure 4C).

In TB-COVID-19 co-infected patients, several biochemical parameters exhibited significant negative correlations. Notably, platelet levels were negatively correlated with hematocrit, and total bilirubin was negatively correlated with D-dimer. Hemoglobin levels were negatively correlated with ALT, and direct bilirubin was negatively correlated with hematocrit and ALT. In contrast, positive correlations were observed between D-dimer and WBC count and between ALT and direct bilirubin levels. Cytokine levels in TB-COVID-19 co-infected patients also showed notable associations. IL-2 levels were negatively correlated with hematocrit, and IL-6 was negatively correlated with both ALT and hematocrit. Urea levels were negatively correlated with IL-4 and IL-6. Lymphocyte count was positively correlated with TNF-α, and D-dimer levels were positively correlated with TNF-α. Additionally, both IL-6 and IL-4 were negatively correlated with urea levels. Several *ace2* and *tmprss2* SNPs demonstrated significant associations with biochemical markers in TB-COVID-19 co-infected patients. Specifically, rs4646142 (an *ace2* SNP) and rs12329760 (a *tmprss2* SNP) were negatively correlated with AST levels. Rs2074192 was negatively correlated with IL-2 but positively correlated with hematocrit. Rs12329760 also showed a negative correlation with WBC count and AST, while rs4646142 exhibited a negative correlation with AST. Rs2285666 was negatively correlated with lymphocyte count, and rs4646140 was positively correlated with direct bilirubin, hematocrit, and ALT levels (Figure 4D).

## Discussion

It has been documented that kidney and liver function could be altered during COVID-19 and TB infections through increased AST, ALT, urea, creatinine and bilirubin levels. In this study, increased serum AST, urea and D-dimer levels were associated with TB-COVID-19 co-infection. Significantly higher serum AST levels observed in the TB-COVID-19 patients compared to this study corroborated earlier reports (18–20). High AST levels in disease groups could be associated with worse outcomes (21). Higher serum urea levels in TB-COVID-19 co-infected patients in this study corroborated with a previous report (22). Increased D-dimer levels in patients with concurrent TB and COVID-19 in this study may indicate elevated thrombotic risks. This is consistent with findings reported in Pakistani populations (23). The elevation of D-dimer, a fibrin degradation product, often correlates with inflammation and coagulopathy, which are critical considerations in managing patients with these comorbidities (24). Monitoring D-dimer levels could be essential for assessing thrombotic risk and guiding treatment strategies in such cases. Elevated serum ALT levels in COVID-19 patients can indeed suggest underlying liver dysfunction, which has been observed in various studies. COVID-19 has been associated with hepatic impairment, and increased ALT is often a marker of liver injury. Zhang et al. noted similar findings, highlighting the importance of monitoring liver function in COVID-19 patients (25).

Given the fact that TB and COVID-19 share similar signs and symptoms, misdiagnosis of disease due to clinical parameters alone is likely to be prevalent. All participants from the diseased group in this study had a high frequency of cough, fever, headache and fatigue, consistent to earlier reports (26–28). One of the major findings reported in this study was an exacerbated immune response in TB-COVID-19 co-infected patients demonstrated by high levels of IFN-γ, TNF-α and IL-10. IFNγ’s role is partially reflected by the fact that it increases the production of proinflammatory cytokines via activation of the JAK-STAT pathway, leading to clinical manifestations of disease (29). Elevated IFN-γ levels in TB-COVID-19 may be contributing to the abnormal systemic inflammatory responses that increase disease severity. While IFN-γ production shows subject-specific variations, reduced levels were reported in active TB patients (25,30). Also, TNF-α, which is a key cytokine involved in inflammatory responses in TB and COVID-19, demonstrated higher levels in TB-COVID-19 comorbid patients. A recent study in mice models found that the combination of TNF-α and IFNγ induced a cytokine-mediated inflammatory cell death signalling pathway via JAK-STAT1 (31). Other pro-inflammatory cytokines such as IL-6, IL-2 and IL-1β showed varying expression levels among the diseased subjects. Higher plasma IL-6 levels observed in TB patients discriminated efficiently from the other study subjects (32–34). IL-6 levels associated with pro-inflammatory cytokines variants contribute to the cytokine storm, deteriorating COVID-19 outcomes (35,36). The higher levels of IL-2 expressed during COVID-19 in this study corroborate other studies (37–40). A previous report demonstrated that IL-2 levels were useful in determining the prognosis of lung damage in influenza A patients (41) .IL-1β, another pro-inflammatory cytokine, was expressed at higher levels in the diseased group compared to the controls. This finding aligns with previous research on TB and COVID-19 patients (42,43) .IL-10, an anti-inflammatory cytokine, was found at significantly higher levels in TB-COVID-19 patients than other groups. Previous studies have also reported elevated IL-10 levels in TB-COVID-19 patients compared to those with either TB or COVID-19 alone, regardless of severity (22,23). Plasma IL-4 levels in TB-COVID-19 co-infected patients were significantly higher than the other diseased and healthy control groups and corroborated earlier report (44).

Despite the availability of data regarding variations in genotype frequencies of *ace2* and *tmprss2* during COVID-19 infection, limited information is available about their implication in TB and COVID-19 co-infections. Our data adds considerable insight to the literature on mutations of *ace2* and *tmprss2* involved in TB and COVID-19 pathogenesis. The present study sought to identify genotypic variations in human *ace2* and *tmprss2* genes and correlate these with susceptibility to these co-infections. Numerous reports demonstrated host *ace2* as the main receptor for SARS-CoV2 and *tmprss2* as responsible for spike protein (3,20,45).

Specific changes in the *ace2* and *tmprss2* gene sequences, which may increase the binding affinity or increased expression, may affect the entry of the SARS-CoV-2 (46). Populations with specific SNPs in the *ace2* and *tmprss2* genes were reported to have/show increased susceptibility to COVID-19 and TB-COVID-19 co-infection (47–49). Such reports on the structural and regulatory variants of *ace2* and *tmprss2* conferring susceptibility to COVID-19 from the Cameroonian population are limited. Reports showed that African populations are genetically predisposed to low *ace2* and *tmprss2* expression, partly explaining the lower incidence of COVID-19 (21). Whereas allelic frequencies contributing to higher *ace2* and *tmprss2* expressions in South Asian, Southeast Asian, and East Asian populations reported higher infection rates (50,51). In this study, we monitored polymorphism patterns in *ace2* and *tmprss2* to find a correlation with higher genetic susceptibility to COVID-19, if any. The high incidence rate of COVID-19 and TB-COVID-19 co-infected males could be potentially attributed to the presence of the *ace2* gene on the X-chromosome (52). A recent report demonstrated an inverse correlation between *ace2* expression levels and estrogen levels in the SARS-CoV-2 patients (53). Estrogen may contribute to the suppression of *ace2* expression and might partly explain the protective factors in females against COVID-19 (54). The prevalence and risk of COVID-19 have been significantly correlated with the heterozygous mutant genotypes (AG and CA) of *ace2* variants rs4646116, rs2285666, and rs4646179, as reported in the Chinese population (55). Notably, despite the predominance of heterozygous mutant alleles of rs2285666 among the COVID-19 group, a meta-analysis indicates that the rs2285666 GA genotype may confer a protective effect against severe COVID-19 (55). In contrast, studies involving Indian COVID-19 patients have found that the wild genotype of variant rs2285666 is associated with increased disease prevalence (56). The genotype frequencies of *ace2* variants rs4646142, rs35803318, rs4646116 (AG), as well as *tmprss2* variants rs75603675 and rs61735791 (CA), were predominantly found in TB patients. The homozygous mutant alleles ace2 rs2074192 AA frequencies were common in TB patients and TB-COVID-19 co-infected patients. In *tmprss2,* the heterozygous mutant alleles rs12329760 CT were predominant in all disease groups, an observation that supports the hypothesis that these SNPs involved pathogenic activity (57,58). The wild alleles of the *ace2* gene of rs4646140 GG were higher in diseased groups than in healthy controls. In COVID-19 patients, rs2285666 was the most common variant having the CA genotype. The second most frequent SNP were rs4646116 and rs4646179, with patients having the AG genotype having the GG genotype of the COVID-19 group. In TB patients, genotype AG of rs4646142 and rs4646116 and genotype CA of rs2285666, rs75603675, and rs61735791 were the most predominant. Although there was variation in SNP frequency across the study population, SNP rs61735791 was notably related to TB and TB-COVID-19 co-infection.

It is important to acknowledge the limitations of a small sample size, as it can restrict the generalizability of the findings. With a larger cohort, the robustness of these conclusions on the relationships between these SNPs and other research indices could be expanded. The survey of polymorphisms from diverse genetic backgrounds might explain the vulnerability to diseases.

In summary, this study indicates that heterozygous mutant genotypes rs147311723, rs35803318 (AG), rs4240157 (CG) and rs4646179 (AG) in *ace2* and *rs61735791 (CA), rs12329760 (CT)* in *tmprss2* genes may be linked to genetic susceptibility to Tuberculosis and COVID-19 co-infection. These findings highlight the need for a human genetics initiative to understand better the genetic factors influencing COVID-19 susceptibility, which could inform prevention and treatment strategies during the future pandemic.

## Method

### Ethics Statement, Study Subject Recruitment and Classification

This study is approved by Cameroon National Ethical Commitee for Research in Human Health (N° 2020/07/1265/CE/CNERSH/SP) in Yaoundé. This study was carried out in compliance with bio-ethical laws and data protection state as well as following good clinical practice. All study participants recruited presented signs and symptoms of a respiratory disease. These participants were enrolled at the Djoungolo District Hospital, Jamot Hospital, Ekoumdoum Baptist Hospital and Red Cross Hospital in Yaoundé during the COVID-19 pandemic from the period of March 2021 to September 2022. Recruited participants were assessed for medical history, tobacco consumption and TB medication history. We assessed the following variables: i) socio-demographic factors such as age, sex, occupation, number of co-inhabitants ii) COVID-19 and TB history, asthma history, HIV status, hypertension, diabetes mellitus, tobacco and alcohol consumption, iii) clinical characteristics such as cough, fever, headache, sore throat, asthenia, chest pain, loss of smell. Demographic details and clinical characteristics are presented in Table 1. Every participant underwent clinical examination and laboratory assessment for COVID-19 and TB. A group of asymptomatic individuals who underwent COVID-19 RDT and TB microscopy and full diagnostics of other infectious diseases that are endemic in our region such as malaria, HIV and Hepatitis were enrolled as healthy controls. Nasopharyngeal samples which tested positive by RDT were considered as COVID-19 positive and sputum microscopy positive subjects were included as active TB patients. Routine laboratory diagnosis of other infectious diseases such as including, malaria, Influenza A and B, HIV and Hepatitis were performed. Exclusion criteria for the study groups were existing comorbidities, co-infections with malaria, Influenza A and B, HIV, Hepatitis and unwillingness to give signed informed consent. Written signed informed consent from every recruit was obtained before the start of the study. Study protocol and consent forms were reviewed and approved by the Centre’s regional ethical committee in Yaoundé. We later classified our study population into four groups: COVID-19, TB, TB-COVID-19 co-infection and a set of healthy controls.

### Nasopharyngeal sample collection and processing

Nasopharyngeal samples were collected from the participants by inserting the swab provided about 2 - 2.5 cm into the nostrils. The HIGHTOP Antigen Rapid Test device manufactured by Qingdao Hightop Biotech Company was used according to manufacturer’s instruction. The QIAamp viral RNA mini kit was used to extract coronavirus RNA from nasopharyngeal samples. A confirmatory Covid 19 diagnosis was later done using real time PCR using the Logix Smart ABC (Cat #: ABC-K-001) test utilizing the patented Co-Primer technology (Satterfield, 2014) according to the manufacturer’s instructions. The Co-Primer triplex assay uses extracted viral RNA to detect Influenza A, B and SARS-CoV-2 (gene RdRp and E-gene) in upper respiratory tract samples and even saliva (Netongo *et al.,* unpublished data).

### Sputum sample collection and processing

Sputum samples were collected using plastic cups 40 mL capacity. After collection, sputum microscopy was carried out. A slide was prepared for each sample, fixed and later stained following the Zeihl-Nelseen staining technique. A confirmatory real-time PCR using the SARAGENE^TM^ *Mycobacterium tuberculosis* test COSARA Diagnostics Ltd India and Logix Smart *Mtb* Kit (Cat #: MTB-K-007)-Co-Diagnostics inc, USA was performed on extracted bacterial DNA according to manufacturers instruction. This test detects the presence or absence of IS6110 and MPB64 genes from *Mycobacterium tuberculosis*. The test kit includes an internal control to identify possible qPCR inhibition and verify the quality of sample extraction.

### Blood sample collection and storage

5ml of whole blood were collected into commercially available anticoagulant-treated tubes (EDTA) and dry tubes. The blood was centrifuged 5000 r.p.m for 10 mins Plasma and serum were obtained, aliquoted and stored at -80°c for future use.

### Evaluation of biochemical markers assays involved in TB and COVID-19 infections

A set of serum biochemical markers AST, ALT, UREA, CREA, BIL-D, BIL-T and D-DIMER. AST, ALT, UREA and CREA were quantified using the PreciseMAX reagent kit according to manufacturer’s instruction and results read on the semi-automatic biochemistry analyzer. Serum levels of bilirubin were determined by the photometric detection of the azo derivatives obtained by the serum reaction with the diazonium ion of sulfamic acid. The measurement of D-dimer in serum was done using the dry fluoroimmunoassay analyzer (WWHS Biotech. Inc Shenzhen, P.R China).

### Measurement of pro-inflammatory and anti-inflammatory cytokine levels involved in COVID-19 and TB

Serum cytokines (IL-6, INF-γ, TNFα, IL-10, IL-2 and IL-1β) and were assayed in serum using sandwich ELISA Origene kits (Origene Technologies, Inc, Rockville, MD 20850, US) according to manufacturer’s instruction.

### DNA extraction and Single nucleotide polymorphism genotyping

Genomic DNA was harvested from the peripheral blood using the commercially available Quick-DNA™ Miniprep Kit (QIAamp DNA Blood Mini kit, Qiagen, Germany), according to manufacturer’s instruction and its quality was verified in agarose gels stained with ethidium bromide nucleic acid gel stain (Thermo Fisher Scientific, C.A, USA). Then, the DNA concentration was measured using nanodrop (Thermo Fisher Scientific, C.A, USAe) and purity was determined by calculating the A260/280 ratio . The extracted genomic DNA were stored at -80°C until used in genotyping reaction. SNPs were analyzed by real-time polymerase chain allelic discrimination technology using TaqMan SNP genotyping assay kit (Thermo Fisher Scientific, Waltham, MA, United States) on a Co-Dx Box Magnetic Induction Cycler qualitative Time Polymerase Chain Reaction (qPCR) machine (Co-Diagnostics Inc USA, Cat # MIC001355)The ten variants analysed for the Angiotensin-converting enzyme gene (ACE 2) were rs2285666, rs4240157, rs4646142, rs4646116, rs6632677, rs4646140, rs147311723, rs2074192, rs35803318, rs4646179 and while three genotype variants were determined for the Transmembrane serine protease 2 Polymorphisms (TMPRSS2) gene,namely rs12329760, rs75603675 and rs61735791. Specifically, genotype variants were determined using the TaqMan™ SNP Genotyping Master Mix kit, from Thermo Fisher Scientific, C.A, USA (Cat #: 4381656) that reveals ACE 2 rs4646179 A>G, rs147311723 G>A, rs4646142 G>A, rs2074192 C>T, rs35803318 C>T, rs4646140 C>T, rs6632677 G>C, rs4646116 T>C, rs2285666 C>A, rs4240157 C>G and TMPRSS2 rs12329760 C>G, rs75603675 C>A, rs61735791 C>A mutations.

The reaction mix of each sample is composed of 5 µL of 2X TaqMan Genotyping Master Mix, 0.5 µL of TaqMan assay (20X), and 4.5 µL RNase free water. The thermal cycling protocol is optimized as follows 950° C for 10 min for AmpliTaq Gold, UP Enzyme Activation, followed by denaturation step at 950° C for 15 s and annealing/extension at 600° C for 1 min for 40 cycles. The qPCR was performed on Co-Diagnostics PCR instrument (Co-Diagnostic, INC, Salt Lake City, USA) and the results were analyzed using Co-Diagnostic Genotyper softwaree. This software was used to plot the findings of the allelic discrimination data as a scatter plot of Allele 1 (VIC® dye) versus Allele 2 (FAM™ dye). Each well of the 96-well reaction plate was represented as an individual point on the plot.

## Data Availability

All data produced in the present work are contained in the manuscript.

## Data analysis and management

The data was anonymized prior to analysis with numerical variables and summarized an and standard deviation. All comparisons of categorical variables cytokines and biochemical biomarkers data were analyzed using Graph Pad Prism 8.0 statistical packages of categorical variables was performed using Chi-square test at significance level (P<0.005, two sided). Arithmetic means and standard deviation were used. Test techniques used include independent student t-test and Oneway ANOVA test, one for comparing 2-independent groups and the other for more than 2-independent groups. SNP frequencies were expressed as numbers (%) in each group, and the Chi-square test was used to analyze the results.

## Ethical Approval

Approval was obtained from the Cameroon National Ethical Committee for Research in Human Health (N° 2020/07/1265/CE/CNERSH/SP) in Yaoundé.

## Consent

Informed consent was obtained from all participants included in the study. All those who refused to sign the written inform consent were excluded from this study.

## Authors’ contributions

Conceptualization: Palmer Masumbe Netongo, Mary Ngongang Kameni, Eric Berenger Tchoupe.

Data curation: Mary Ngongang Kameni, Eric Berenger Tchoupe.

Formal analysis: Mary Ngongang Kameni, Nikhil Bhalla, Ranjan Kumar Nanda Severin Donald Kamdem.

Funding acquisition: Palmer Masumbe Netongo and John Amuasi.

Investigation: Mary Ngongang Kameni, Eric Berenger Tchoupe, Nikhil Bhalla, Tepa Arnaud, Fuh Roger Neba.

Methodology: Mary Ngongang Kameni, Eric Berenger Tchoupe, Tepa Arnaud, Fuh Roger Neba, Severin Donald Kamdem, Palmer Masumbe Netongo.

Project administration: Palmer Masumbe Netongo, Ranjan Kumar Nanda, Assam Assam Jean Paul Anthony Afum-Adjei Awuah, and John Amuasi.

Resources: Palmer Masumbe Netongo, Anthony Afum-Adjei Awuah and John Amuasi. Supervision: Palmer Masumbe Netongo, Ranjan Kumar Nanda, Assam Assam Jean Paul The first draft was prepared by Mary Ngongang Kameni.

Writing – review & editing: Mary Ngongang Kameni, Nikhil Bhalla, Eric Berenger Tchoupe, Severin Donald Kamdem, Ranjan Kumar Nanda, Palmer Masumbe Netongo.

Validation: Mary Ngongang Kameni, Eric Berenger Tchoupe, Severin Donald Kamdem, Nikhil Bhalla, Ranjan Kumar Nanda, Anthony Afum-Adjei Awuah, John Amuasi and Palmer Masumbe Netongo.

