## Supplementary document for "Human *ace2* and *tmprss2* polymorphisms for predicting susceptibility to tuberculosis and COVID-19 co-infection in Cameroonian cohort"

**Supplementary Table 1: Genotype frequencies distribution of single nucleotide polymorphisms of *ace2* and *tmprss2* genes.** Genomic DNA isolated using the Zymo Human DNA Isolation Kit was subjected to quantitative real-time PCR with TaqMan probes. TaqMan SNP Genotyping Assays were employed to identify SNPs in *ace2* and *tmprss2* genes. Chi-square test was used to determine statistically significant differences in genotype frequencies among the groups (Control, COV, TB, and TBCOV). A *p-value* of less than 0.05 was considered significant. Abbreviations: TB: Tuberculosis positive; COV: COVID-19 positive and TBCOV: Tuberculosis and COVID-19 co-infection.

|  | | **Groups** | **Genotypes** | | | **P-value** |
| --- | --- | --- | --- | --- | --- | --- |
|  |  |  | **Homozygous GG** | **Heterozygous AG** | **Homozygous AA** |  |
| ***ace2*** | **rs4646142** | Control (n= 24) | 2 | 22 | 0 | 0.03 |
|  |  | COV (n = 31) | 6 | 25 | 0 |  |
|  |  | TB (n = 43) | 0 | 43 | 0 |  |
|  |  | TBCOV (n = 21) | 3 | 17 | 1 |  |
|  | **rs2074192** | Control (n= 24) | 4 | 20 | 0 | <0.05 |
|  |  | COV (n = 31) | 7 | 24 | 0 |  |
|  |  | TB (n = 43) | 0 | 9 | 32 |  |
|  |  | TBCOV (n = 21) | 2 | 3 | 17 |  |
|  | **rs147311723** | Control (n= 24) | 0 | 24 | 0 | <0.05 |
|  |  | COV (n = 31) | 8 | 18 | 5 |  |
|  |  | TB (n = 43) | 3 | 29 | 11 |  |
|  |  | TBCOV (n = 21) | 1 | 20 | 0 |  |
|  | **rs35803318** | Control (n= 24) | 1 | 23 | 0 | ns |
|  |  | COV (n = 31) | 6 | 25 | 0 |  |
|  |  | TB (n = 43) | 1 | 42 | 0 |  |
|  |  | TBCOV (n = 21) | 0 | 21 | 0 |  |
|  | **rs4646140** | Control (n= 24) | 8 | 12 | 4 | 0.04 |
|  |  | COV (n = 31) | 19 | 10 | 2 |  |
|  |  | TB (n = 43) | 22 | 21 | 0 |  |
|  |  | TBCOV (n = 21) | 13 | 5 | 3 |  |
|  | **rs4646116** | Control (n= 24) | 0 | 23 | 1 | <0.05 |
|  |  | COV (n = 31) | 1 | 30 | 0 |  |
|  |  | TB (n = 43) | 0 | 43 | 0 |  |
|  |  | TBCOV (n = 21) | 2 | 18 | 1 |  |
|  |  |  | Homozygous CC | Heterozygous CG | Homozygous GG |  |
|  | **rs4240157** | Control (n= 24) | 3 | 21 | 0 | ns |
|  |  | COV (n = 31) | 3 | 28 | 0 |  |
|  |  | TB (n = 43) | 13 | 30 | 0 |  |
|  |  | TBCOV (n = 21) | 2 | 19 | 0 |  |
|  | **rs6632677** | Control (n= 24) | 0 | 20 | 4 | 0.05 |
|  |  | COV (n = 31) | 1 | 29 | 1 |  |
|  |  | TB (n = 43) | 0 | 40 | 3 |  |
|  |  | TBCOV (n = 21) | 1 | 14 | 6 |  |
|  |  |  | Homozygous CC | Heterozygous CA | Homozygous GG |  |
|  | **rs2285666** | Control (n= 24) | 0 | 24 | 0 | 0.004 |
|  |  | COV (n = 31) | 0 | 31 | 0 |  |
|  |  | TB (n = 43) | 0 | 43 | 0 |  |
|  |  | TBCOV (n = 21) | 3 | 17 | 1 |  |
|  |  |  | Homozygous AA | Heterozygous AG | Homozygous GG |  |
|  | **rs4646179** | Control (n= 24) | 0 | 24 | 0 | ns |
|  |  | COV (n = 31) | 1 | 30 | 0 |  |
|  |  | TB (n = 43) | 2 | 40 | 1 |  |
|  |  | TBCOV (n = 21) | 2 | 19 | 0 |  |
| ***tmprss2*** |  |  | Homozygous CC | Heterozygous CA | Homozygous AA |  |
|  | **rs75603675** | Control (n= 24) | 0 | 24 | 0 | ns |
|  |  | COV (n= 31) | 1 | 29 | 1 |  |
|  |  | TB (n = 43) | 0 | 43 | 0 |  |
|  |  | TBCOV (n= 21) | 1 | 18 | 2 |  |
|  |  |  | Homozygous CC | Heterozygous CA | Homozygous AA |  |
|  | **rs61735791** | Control (n= 24) | 0 | 24 | 0 | ns |
|  |  | COV (n= 31) | 3 | 28 | 0 |  |
|  |  | TB (n = 43) | 0 | 43 | 0 |  |
|  |  | TBCOV (n= 21) | 0 | 21 | 0 |  |
|  |  |  | Homozygous CC | Heterozygous CT | Homozygous TT |  |
|  | **rs12329760** | Control (n= 24) | 2 | 22 | 0 | ns |
|  |  | COV (n= 31) | 2 | 29 | 0 |  |
|  |  | TB (n= 43) | 5 | 38 | 0 |  |
|  |  | TBCOV (n=21) | 2 | 19 | 0 |  |

***End of Supplementary Document***
